# Do people with low back pain walk differently? A systematic review and meta-analysis

**DOI:** 10.1101/2021.05.08.21256890

**Authors:** Jo Armour Smith, Heidi Stabbert, Jennifer J. Bagwell, Hsiang-Ling Teng, Vernie Wade, Szu-Ping Lee

## Abstract

**Objective:** To identify differences in biomechanics during gait in individuals with acute and persistent low back pain compared with back-healthy controls.

**Design:** Systematic review

**Data Sources:** A search was conducted in PubMed, CINAHL, SPORTDiscus, and PsycINFO in June 2019 and was repeated in December 2020.

**Eligibility criteria:** Studies were included if they reported biomechanical characteristics of individuals with and without low back pain during steady-state or perturbed walking and running. Biomechanical data included spatiotemporal, kinematic, kinetic, and electromyography variables. The reporting quality and potential for bias of each study was assessed. Data were pooled where possible to compare the standardized mean differences (SMD) between groups.

**Results:** Ninety-seven studies were included. Two studies investigated acute pain and the rest investigated persistent pain. Eight studies investigated running gait. 20% of studies had high reporting quality/low risk of bias. In comparison with back-healthy controls, individuals with persistent low back pain walked more slowly (SMD -0.59 [95% CI -0.77 to -0.42]) and with shorter stride length (−0.38 [-0.60 to -0.16]). There were no differences in the amplitude of motion in the thoracic or lumbar spine, pelvis, or hips in individuals with LBP. During walking, coordination of motion between the thorax and the lumbar spine/pelvis was significantly more in-phase in the LBP groups (−0.60 [-0.90 to -0-.30]), and individuals with LBP exhibited greater amplitude of activation in the paraspinal muscles (0.52 [0.23 to 0.80]).

**Summary/Conclusion:** There is moderate to strong evidence that individuals with persistent LBP demonstrate impairments in walking gait compared with back-healthy controls.

## INTRODUCTION

The experience of acute low back pain (LBP) is almost universal, with up to 80% of people experiencing an acute episode of LBP in their lifetimes[1]. However, the greatest burden to individuals and society comes from the pain and disability associated with persistent LBP[2,3]. Persistent LBP is characterized by symptoms lasting or recurring over months and years[4]. Recently, researchers have differentiated between persistent LBP that is experienced almost every day (chronic LBP) and persistent LBP that follows a more episodic pattern (recurrent LBP)[5,6]. Although there are attempts to standardize definitions for recurrent and chronic patterns of persistent LBP[5,7,8], these definitions have not yet been widely adopted.

Walking and running gait are frequently assessed in individuals with acute and persistent LBP during clinical evaluations and as part of observational and interventional research. Biomechanical gait impairments in individuals with LBP may be captured by changes in spatiotemporal characteristics like speed or step length, kinematic characteristics like joint/segmental motion or coordination between joints/segments, kinetic characteristics like forces and torques, and electromyography (EMG) characteristics like amplitude or timing of muscle activation. The amount of trunk motion and joint loading during gait is relatively low[9– 11]. Despite this, due to the repetitive, cyclical nature of walking and running, it is theorized that adverse loading over time in response to changes in gait mechanics in the trunk or lower limbs may contribute to the onset or recurrence of LBP symptoms. Recent work has highlighted the inconsistent evidence for biomechanical impairments during tasks such as gait in individuals with persistent LBP[12]. In part, this inconsistency is due to heterogeneity in clinical back pain presentations[12] and is worsened by small sample sizes in individual studies. In order to develop appropriate rehabilitation strategies for back pain, it is critical to first determine if there are biomechanical impairments during important functional activities such as walking and running that generalize across individuals with LBP. Of the two recent reviews investigating gait in individuals with LBP[13,14], neither performed a quantitative synthesis of the results, and one only included EMG data[14].

The aims of this systematic review therefore were to review and quantitatively synthesize evidence for differences in walking and running gait biomechanics in individuals with acute and persistent low back pain compared with back-healthy controls.

## METHODS

This review was conducted based on the Preferred Reporting Items for Systematic Reviews and Meta-Analyses guidelines (PRISMA). The protocol was registered in the International Prospective Register of Systematic Reviews (PROSPERO: CRD42018078746).

### Search Strategy

The search was conducted in the PubMed, CINAHL, SPORTDiscus, and PsycINFO databases in June 2019, without date restriction. The search terms combined keywords or MeSH terms for gait AND low back pain and were tailored for specific databases. The search strategy is shown in full in the supplementary material. After removal of duplicates, two authors (JAS and VW) double screened title and abstracts based on the inclusion criteria (described in detail below). Full text manuscripts of remaining articles were then retrieved and additionally screened. Reference lists from retrieved articles, previous systematic reviews, and NCBI citation alerts were also checked. In September 2020, the search was repeated using identical search terms in the same databases to identify studies published since the original search.

### Inclusion criteria

Included studies had to be peer reviewed, original research works that were available in English. Eligible studies compared gait variables between a group of individuals with acute or persistent LBP, and a group of back-healthy controls. Study types included case-control, cross-sectional or prospective cohort studies. Studies had to include objectively quantified gait data in one or more of the following categories: 1) spatiotemporal data (speed, distance, step and stride characteristics); 2) kinematic data (peak excursion or total range of motion in the thoracic or lumbar spine, pelvis or lower extremities or coordination in kinematics between two or more joints/segments); 3) kinetic data (net joint moments, joint impulse, work, power); 4) ground reaction force data (vertical or horizontal ground reaction forces); 5) EMG (amplitude or timing of activation in the trunk or lower extremity musculature). Gait paradigms included overground and treadmill steady-state walking and running, as well as walking under dual task conditions involving additional mechanical or cognitive tasks. In studies that included pre- and post-intervention data, only the pre-intervention outcomes were included in this review. Studies were excluded if they were conference abstracts, case reports, dissertations, or review articles, if they did not report comparisons between individuals with and without LBP, or if LBP was experimentally induced in previously asymptomatic participants.

### Study quality assessment

Quality of the reporting of the included studies, and the risk of bias, was assessed using a 16-criteria checklist[15–18] (Table 1). A positive score was given for each criterion met by the study. A total quality score was calculated as the sum of all positive scores from criteria 3 through 16 relevant to the study type (8, 12, and 9 for cross-sectional, case-control, and prospective cohort studies respectively) and a percentage of the possible maximum score was calculated. Each study was independently scored by two authors (JAS scored all studies, S-PL, JB and H-LT scored one third of the studies each). Where there was a difference in scores, the two scoring authors discussed the criteria where the scoring discrepancy occurred and reached a consensus on a final score. Studies were designated as having high reporting quality if they scored 50% or more[17].

**Table 1.** Checklist for assessment methodological quality for cross sectional (CS), case-control (CC) and prospective cohort (PC) study designs.

| Domain | Item # | Description | CS | CC | PC |
| --- | --- | --- | --- | --- | --- |
| Study objective | 1 | Positive, if the study had a clearly defined objective | + | + | + |
| Study population | 2 | Positive, if the main features of the study population are described (sampling frame and distribution of the population according to age and sex) | + | + | + |
|  | 3 | Positive, if cases and controls are drawn from the same population and a clear definition of cases and controls is given and if subjects with the disease/symptom in the past 3 months are excluded from the control group |  | + |  |
|  | 4 | Positive, if the participation rate is at least 80% or if the participation rate is 60–80% and the non-response is not selective (data shown) | + | + | + |
|  | 5 | Positive, if the participation rate at main moment of follow-up is at least 80% or if the non-response is not selective (data shown) |  |  | + |
| Measurements | 6 | Positive, if data on history of the disease/symptom is collected and included in the statistical analysis | + | + | + |
|  | 7 | Positive, if the outcome is measured in an identical manner among cases and controls |  | + |  |
|  | 8 | Positive, if the outcome assessment is blinded with respect to disease status | + | + |  |
|  | 9 | Positive, if the outcome is assessed at a time before the occurrence of the disease/symptom |  | + |  |
| Assessment of the outcome | 10 | Positive, if the time-period on which the assessment of disease/symptom was based was at least 1 year |  |  | + |
|  | 11 | Method for assessing injury status: physical examination blinded to exposure status (+); self-reported: specific questions relating to symptoms/disease/use of manikin (+), single question (–) | + | + | + |
|  | 12 | Positive, if incident cases were included (prospective enrolment) |  | + |  |
| Analysis and data presentation | 13 | Positive, if the measures of association or group comparisons estimated were presented including confidence intervals | + | + | + |
|  | 14 | Positive, if the analysis is controlled for confounding or effect modification: individual factors | + | + | + |
|  | 15 | Positive, if the analysis is controlled for confounding or effect modification: other factors | + | + | + |
|  | 16 | Positive, if the number of cases in the final multivariate model was at least 10 times the number of independent variables in the analysis | + | + | + |
| **Total possible score (sum of items 3 – 16)** |  |  | **8** | **12** | **9** |

### Data extraction

The following data were extracted from all eligible studies: study design, sample size, study inclusion/exclusion criteria, study population demographic characteristics, any additional metrics characterizing the LBP cohort, and the biomechanical outcomes. Data were extracted and double checked by three authors (HS, VW, JAS).

Data were synthesized qualitatively if there were at least two studies/unique cohorts with equivalent outcomes. Data were pooled for meta-analysis for outcomes in which there were equivalent data available from more than three studies/unique cohorts. Where necessary, authors of studies that did not report group means/standard deviations were contacted to provide these data. Group averages/standard deviations were calculated from confidence intervals, standard errors, effect sizes and median/inter-quartile ranges as needed using standard methods. For the pooled analyses, group averages/standard deviations from LBP or male/female subgroups reported in some studies were combined[19]. A random effects model was used to calculate standardized mean differences (SMD) and 95% confidence intervals between the LBP and back-healthy groups (Review Manager version 5.4.1)[20]. Effect sizes of > 0.8 were considered large and > 0.5 were considered moderate. The heterogeneity in the results within the pooled analyses was evaluated for each outcome using the chi-squared test to detect significant heterogeneity, and the I^2^ statistic to quantify the heterogeneity, with I^2^ greater than 0.75 indicative of substantial heterogeneity[21]. Studies were excluded from pooled data analyses if the 95% confidence interval for the group effect in that study did not overlap with the confidence interval for the SMD effect, and the removal of the outlier study did not affect the direction or significance of the pooled effect[21].

The level of evidence for the pooled analyses was defined using the following criteria[22,23]: 1) Strong evidence – homogenous data (Chi^2^ P value ≥ 0.05) pooled from studies of which at least two were high quality; 2) Moderate evidence – either heterogeneous data (Chi^2^ P value < 0.05) pooled from studies of which one was high quality, or homogenous data (Chi^2^ P value ≥ 0.05) from lower quality studies; 3) Limited evidence – heterogeneous data (Chi^2^ P value < 0.05) from lower quality studies[22,23].

## RESULTS

The initial search identified 3,272 articles (Figure 1). Following the removal of duplicates, 2,202 articles were available for further evaluation. An additional seven articles were identified manually and during the repeat search in 2020. The abstracts and titles of 2,209 articles were screened. Lastly, 124 full text articles were retrieved and assessed for inclusion. A total of 97 articles were included. Attempts were made to contact authors of 22 studies that did not present group data for one or more variables of interest, and responses were received for seven studies. Median score for reporting quality/risk of bias was 33% (range, 13 - 89%). Nineteen studies scored greater than 50%. Most studies did not report the participation rate and therefore the potential influence of non-response was unclear. Very few studies reported blinding of researchers or presented confidence intervals in their analyses.

**Figure 1.**
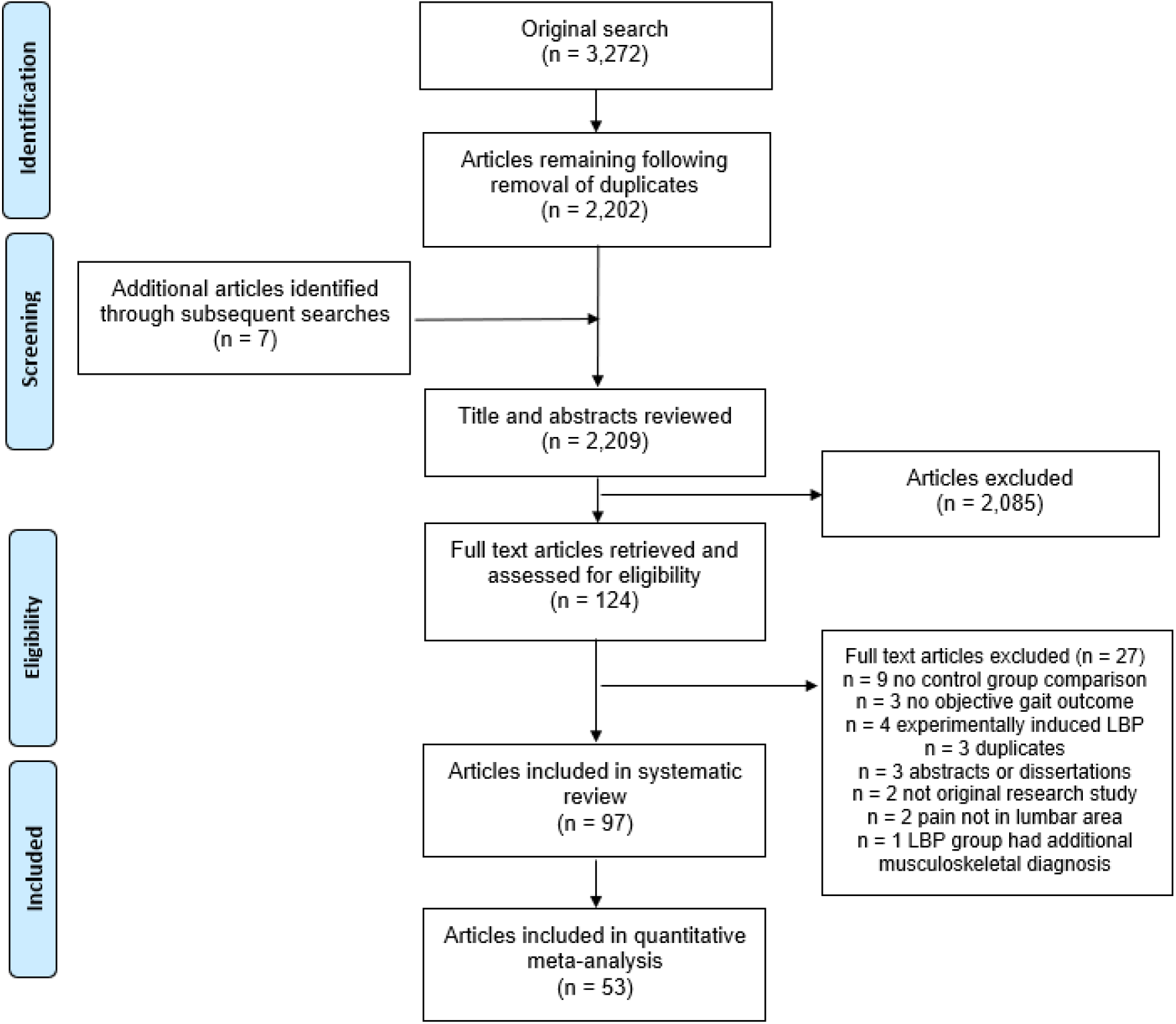
Flow diagram summarizing study selection processes.

### Participant numbers and demographics

Of the included articles, 93 were case-control, 3 were cross-sectional[24–26], and one was a prospective cohort study[27]. Eight studies involving four unique cohorts investigated running gait. Eighty-three studies contained unique sample groups, for a total of, 3,364 individuals with ongoing LBP and 2,315 back-healthy controls. Of the studies that reported participants’ sex, the LBP group included 767 males and 2,075 females and the control group included 653 males and 961 females. The range of mean age for the LBP groups was 21.4 -73.6 years and 18.7-73.5 years for the control groups. Participants with LBP were described as having back pain, nonspecific or idiopathic LBP, chronic LBP, or recurrent LBP. However, as the criteria for these categories varied between studies, we did not attempt to sub-group participants based on these descriptors. The most common measure to define persistent LBP was duration of symptoms, with minimum duration varying from 6 weeks to 1 year, and the most frequent criterion being 3 months (Table 2). Two studies included participants described as having acute LBP, defined as symptom duration of less than seven days, who were re-tested once symptoms had resolved[28,29]. Four studies sharing the same full or partial cohort included a separate group with a history of resolved LBP[30–33]Nineteen studies quantified minimum or maximum pain severity as part of their inclusion criteria (Table 2). Seven studies required LBP to be severe enough to impact function[34–40]. Fourteen studies used pain frequency as a measure to define chronicity or recurrence of episodes[34–38,41–49]. Location of LBP was defined in 16 studies, with location predominantly described as occurring below the costal margin and above gluteal folds (Table 2).

**Table 2.**
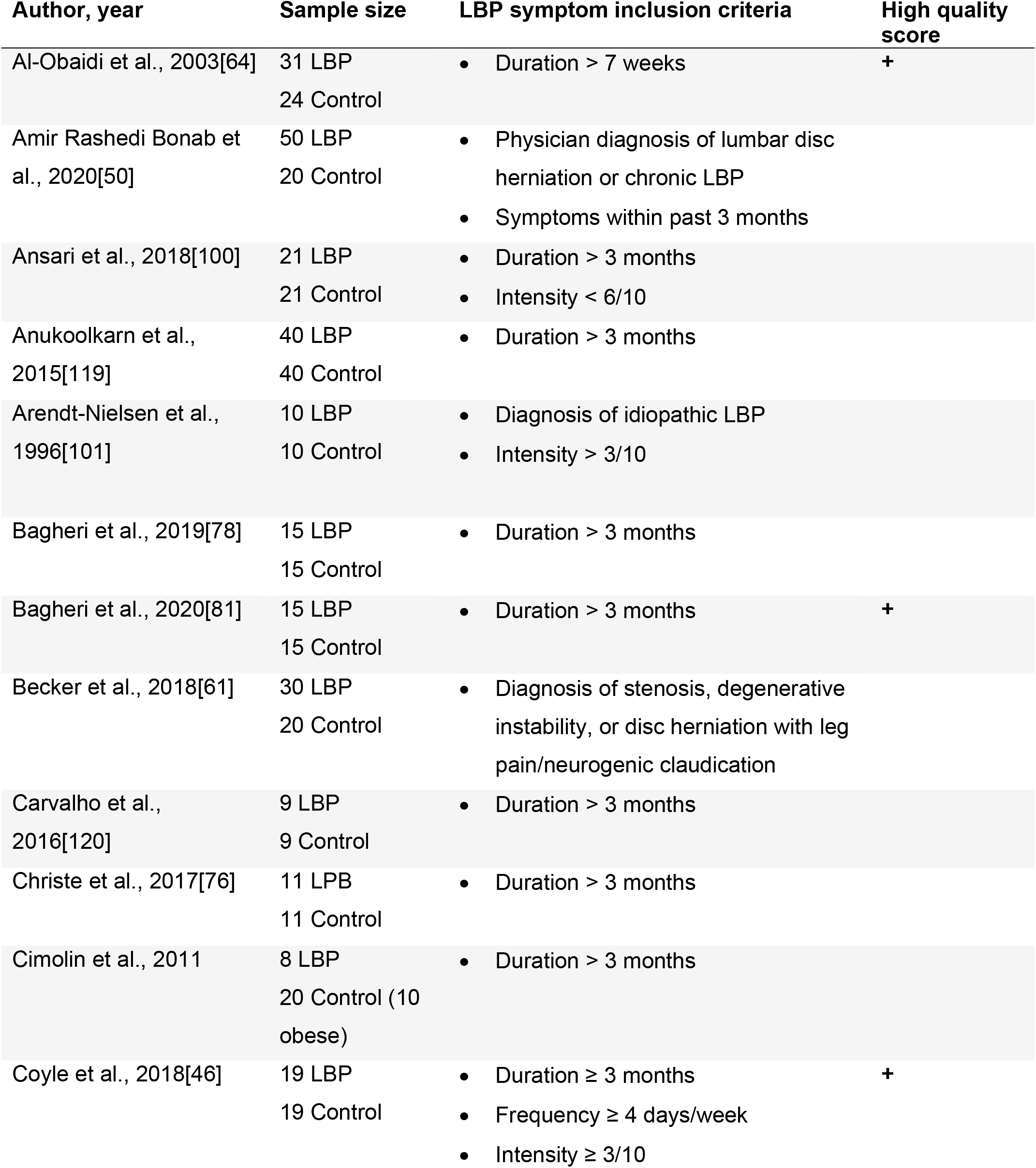
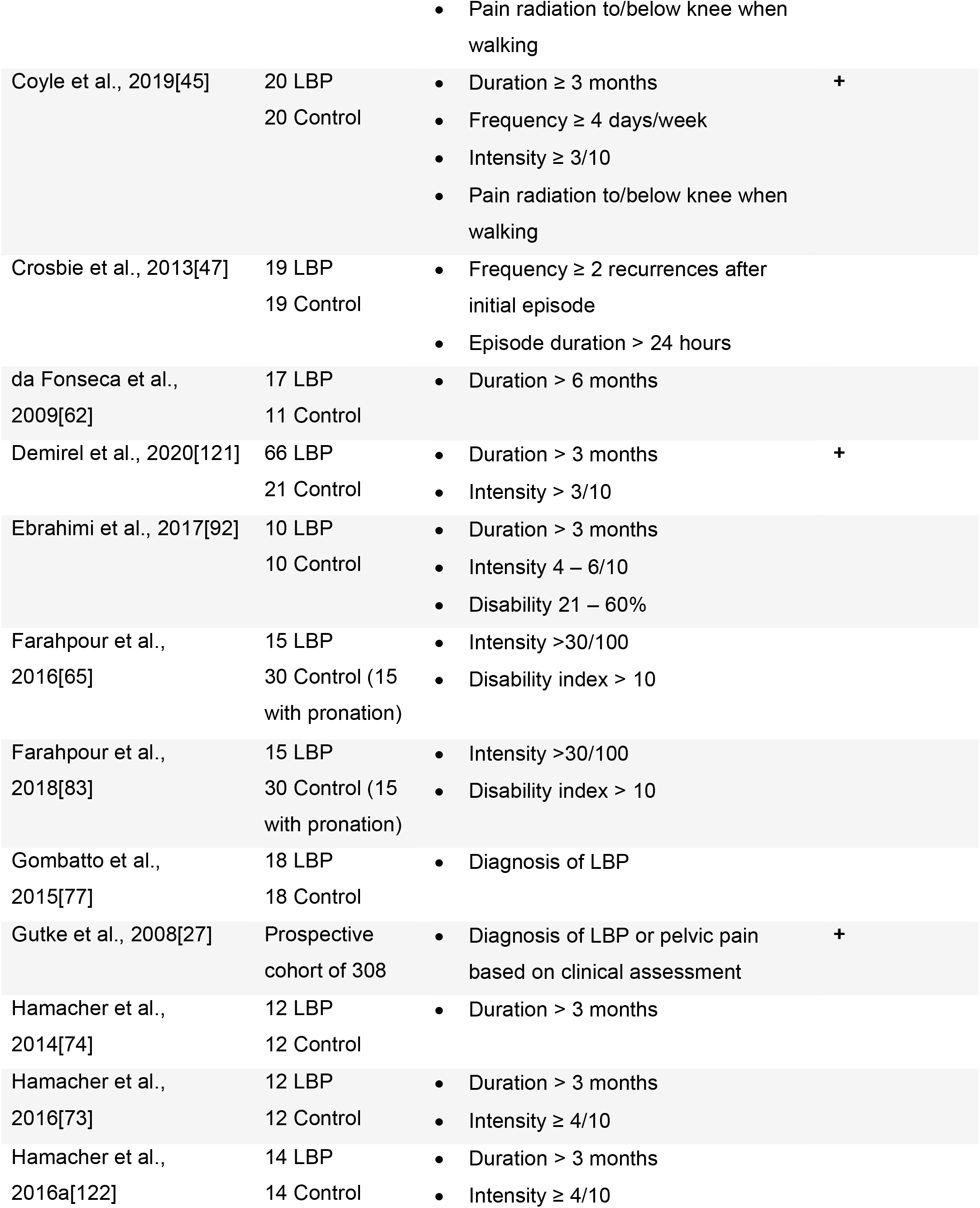
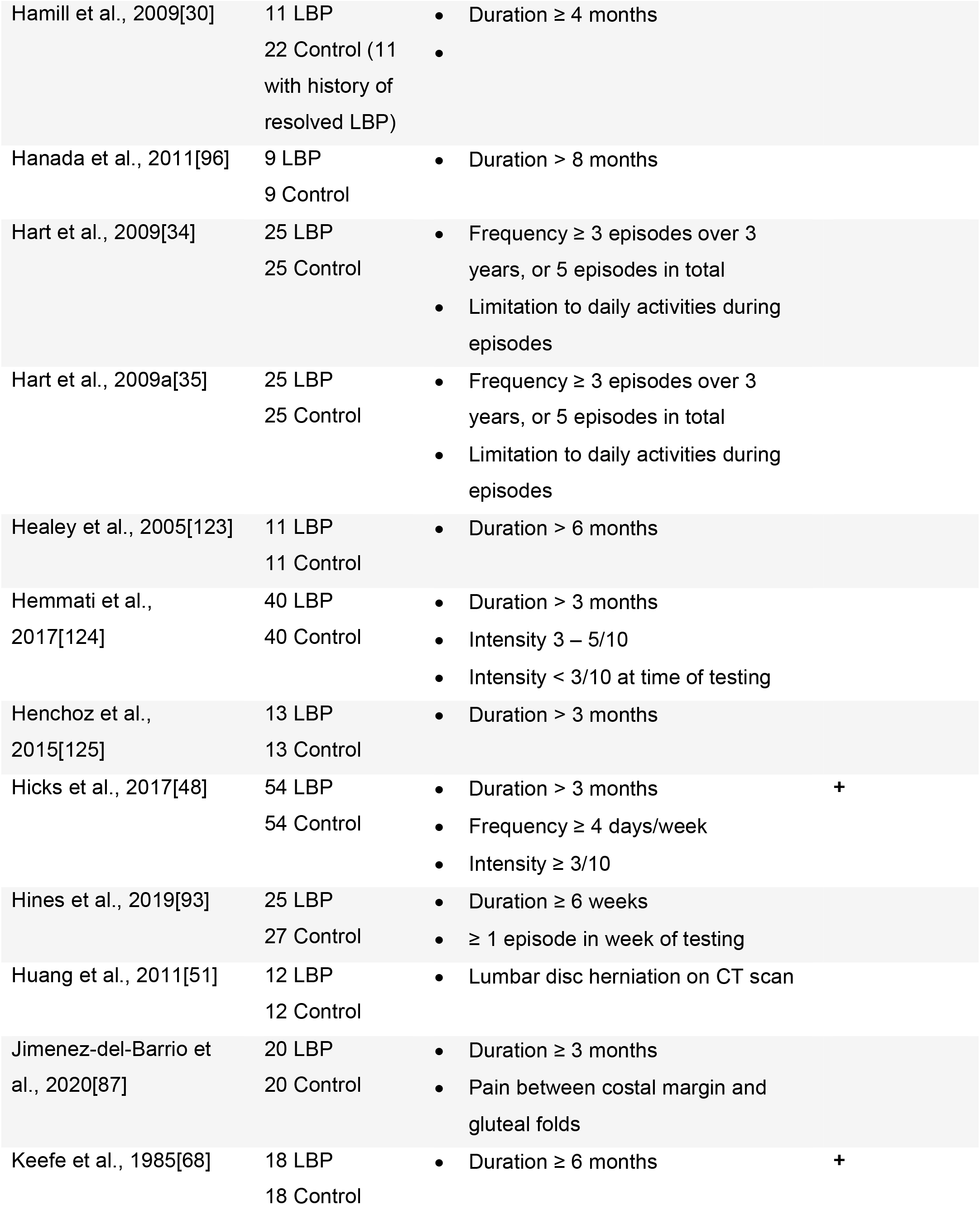
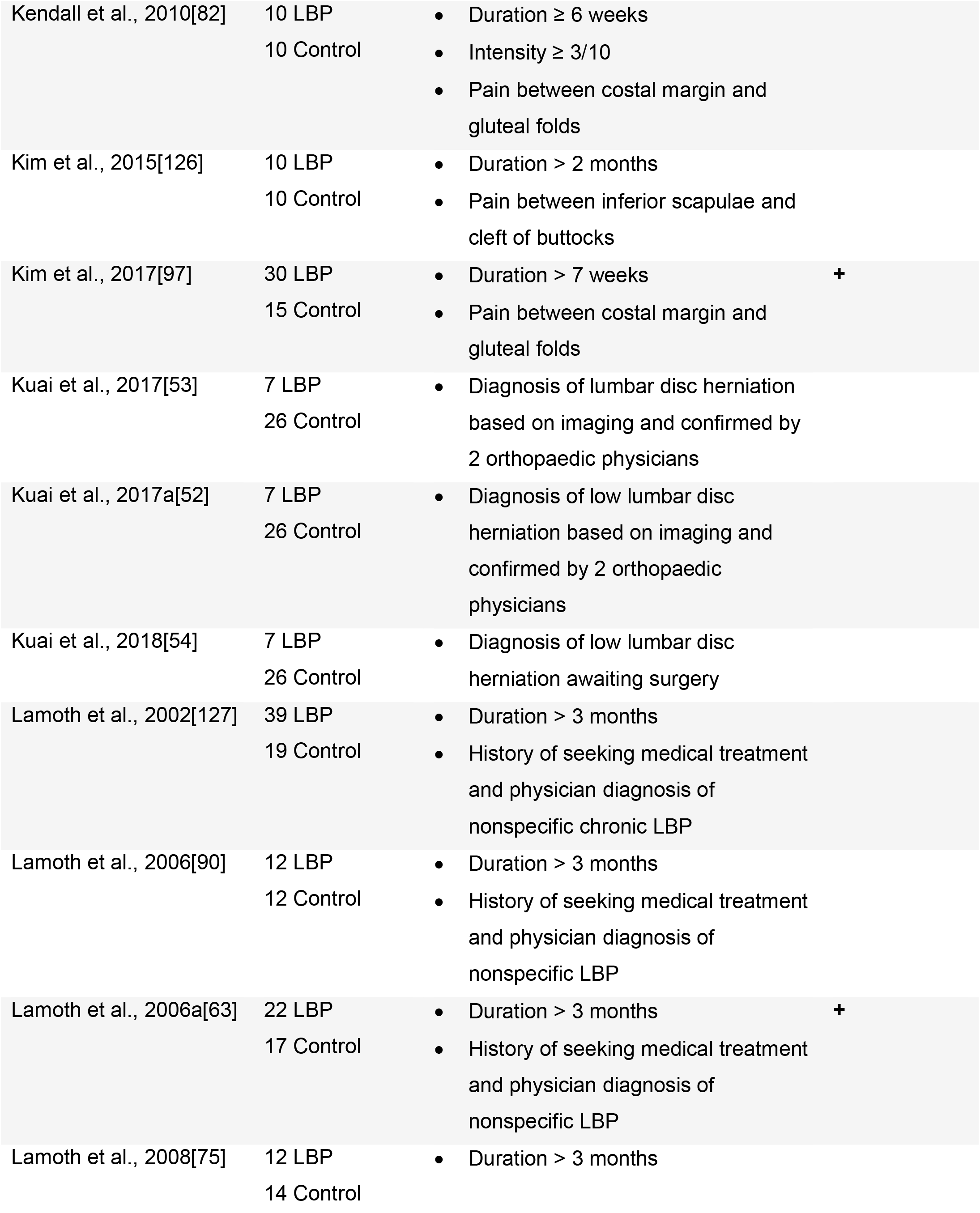
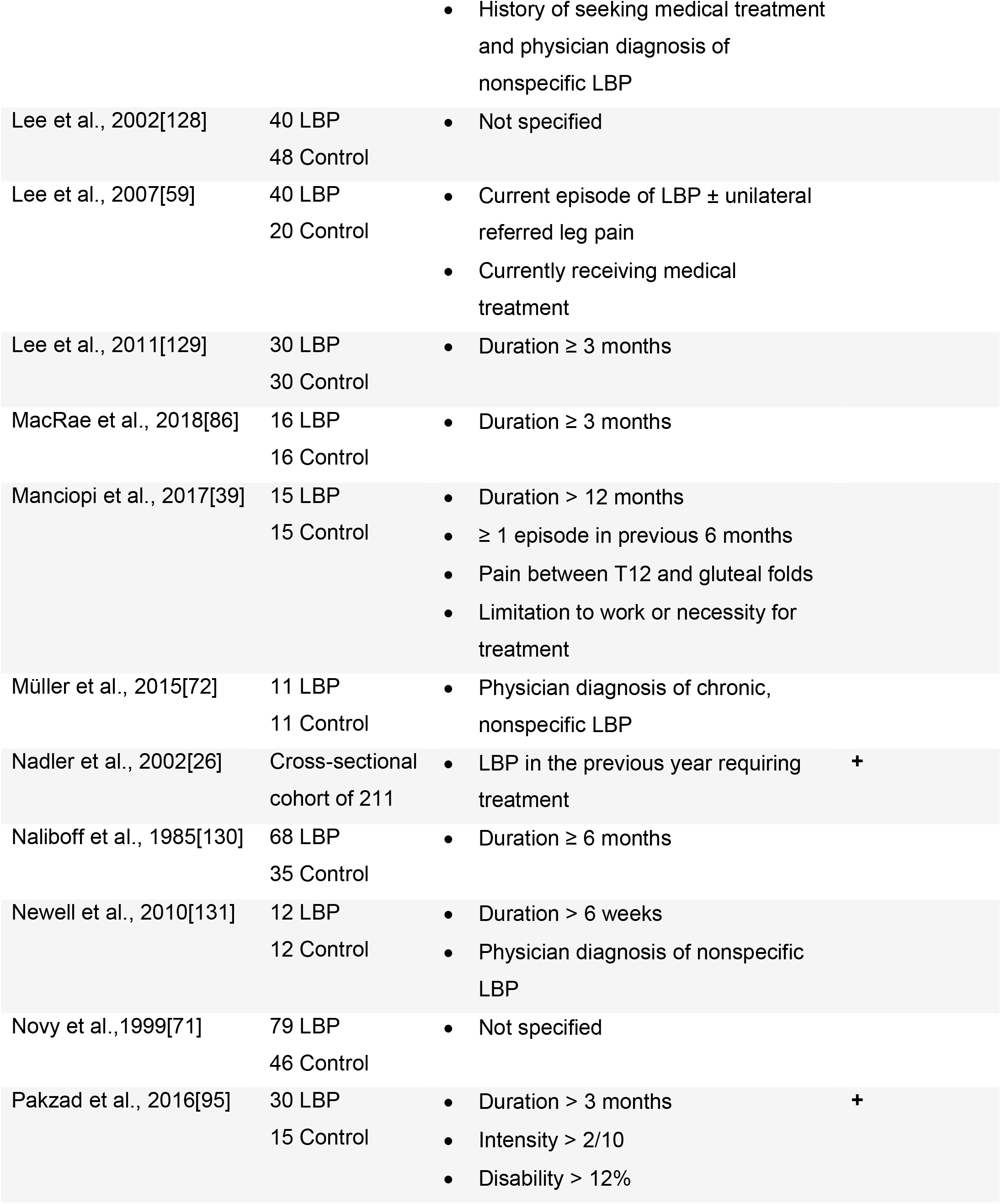
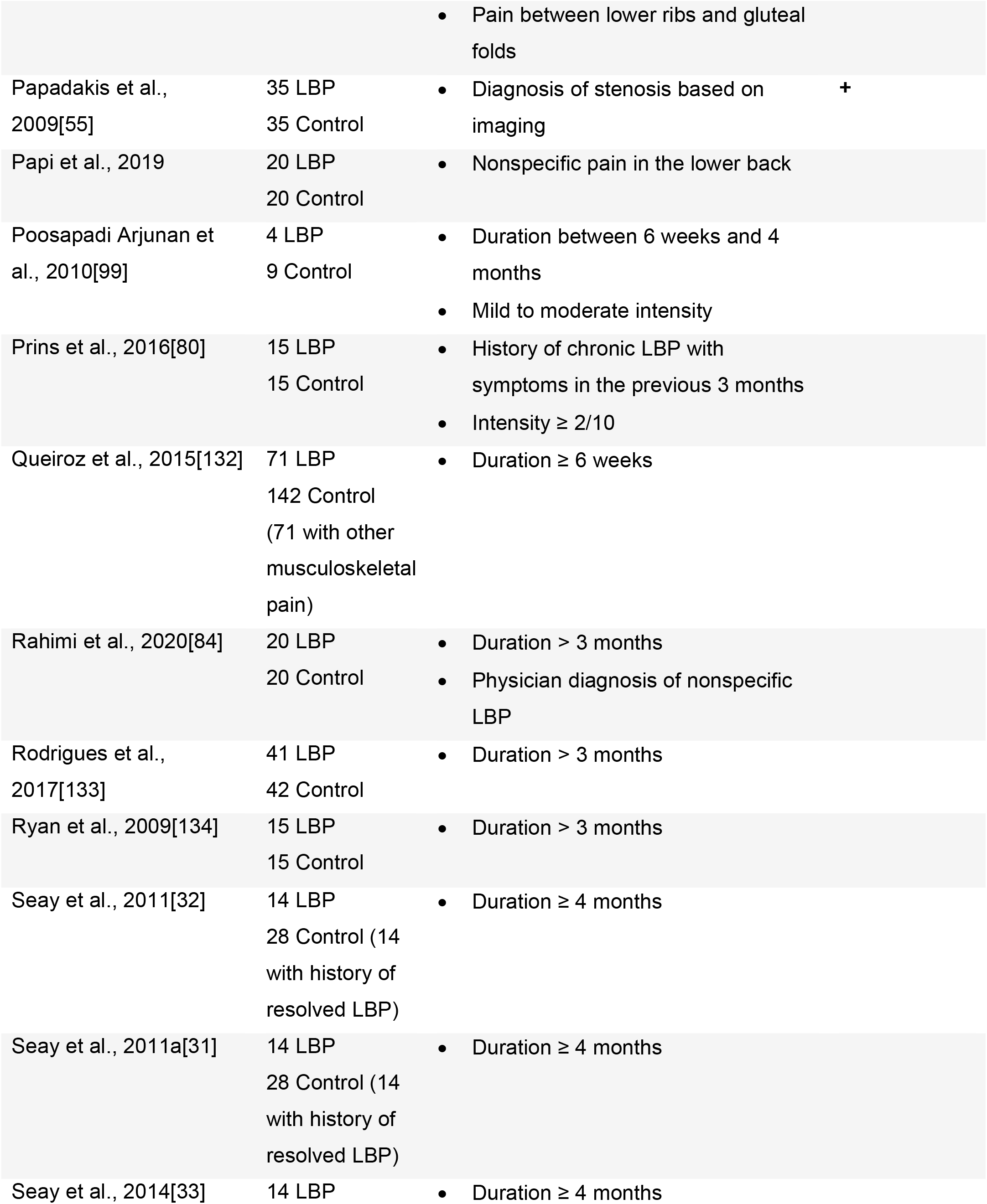
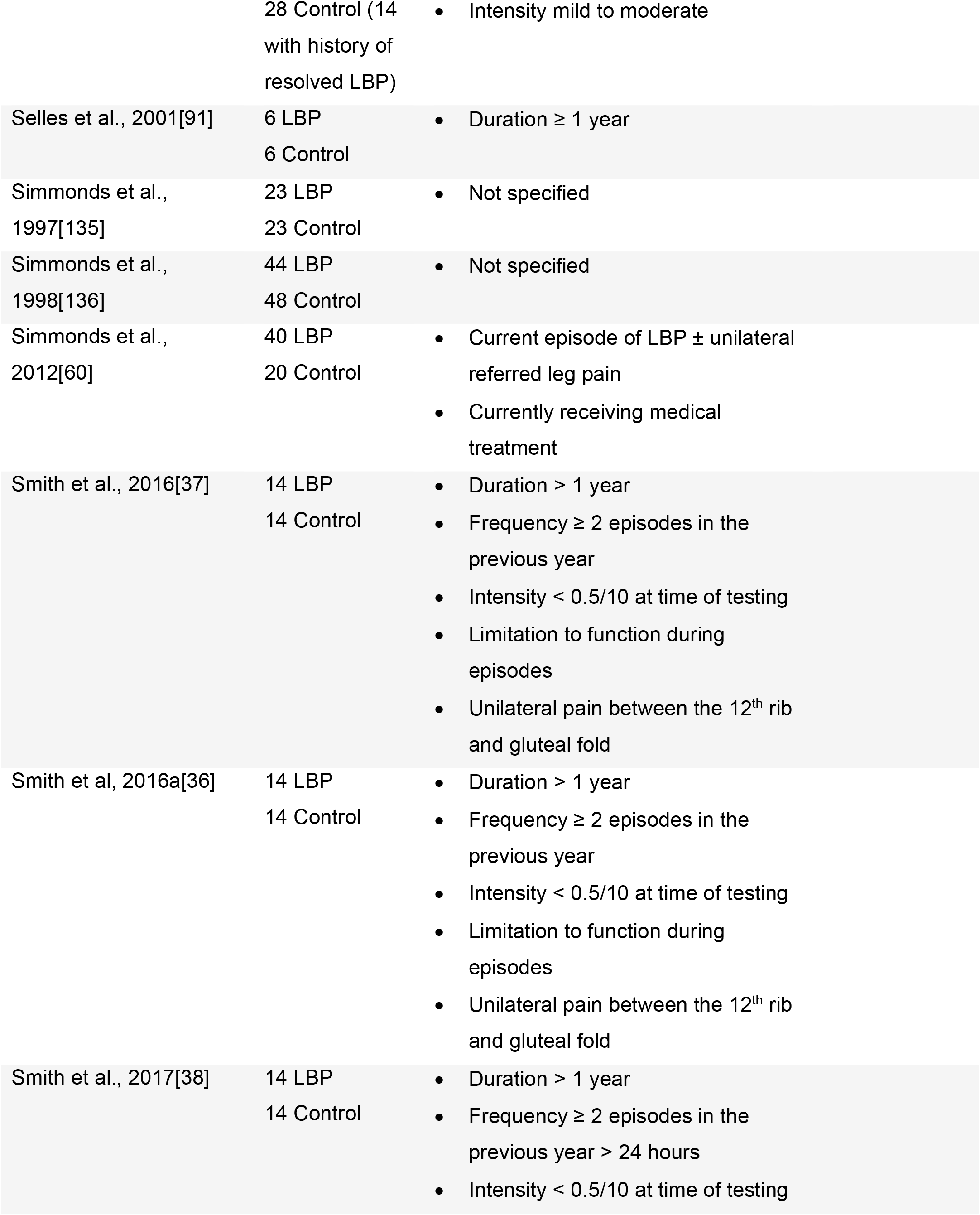
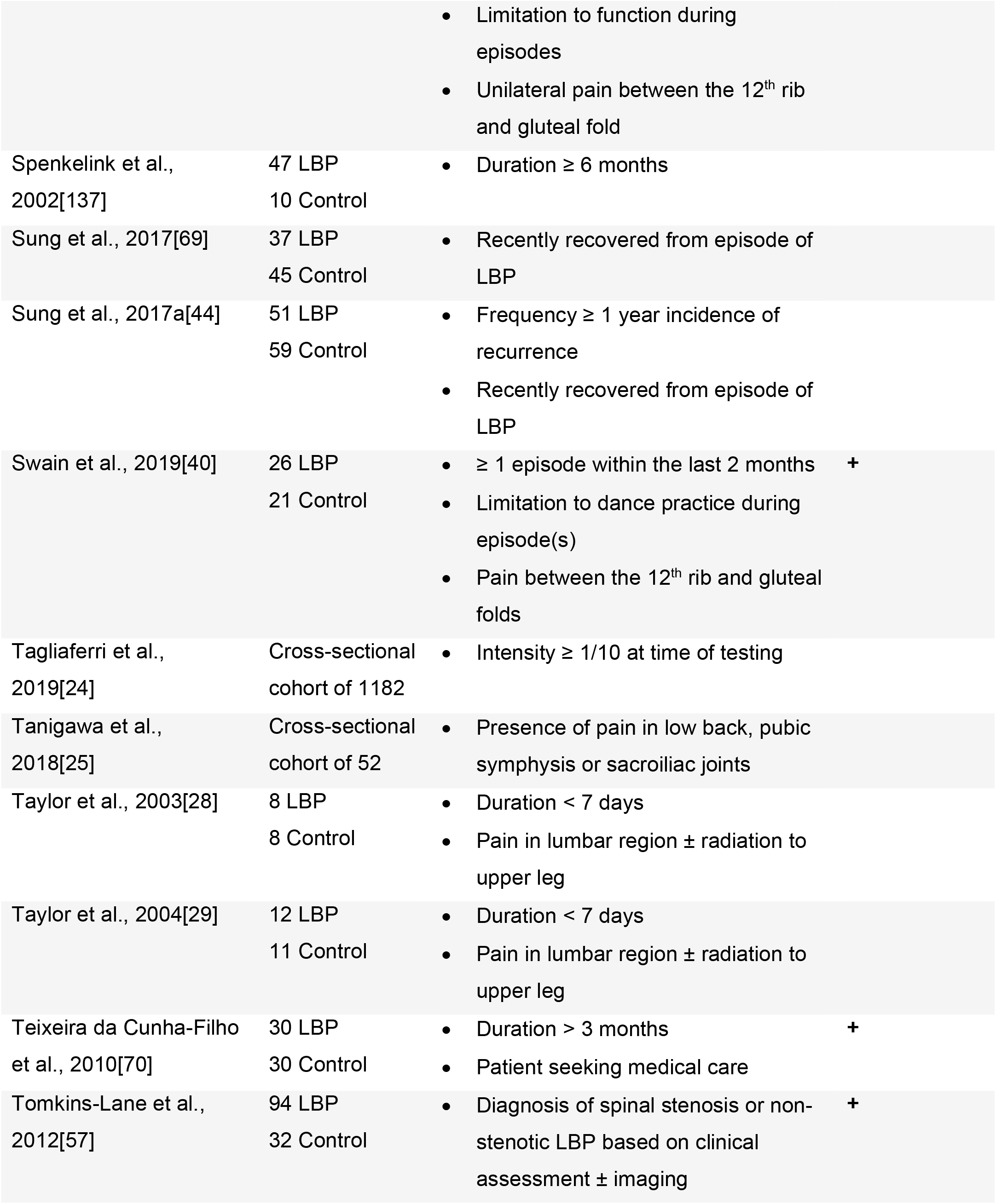
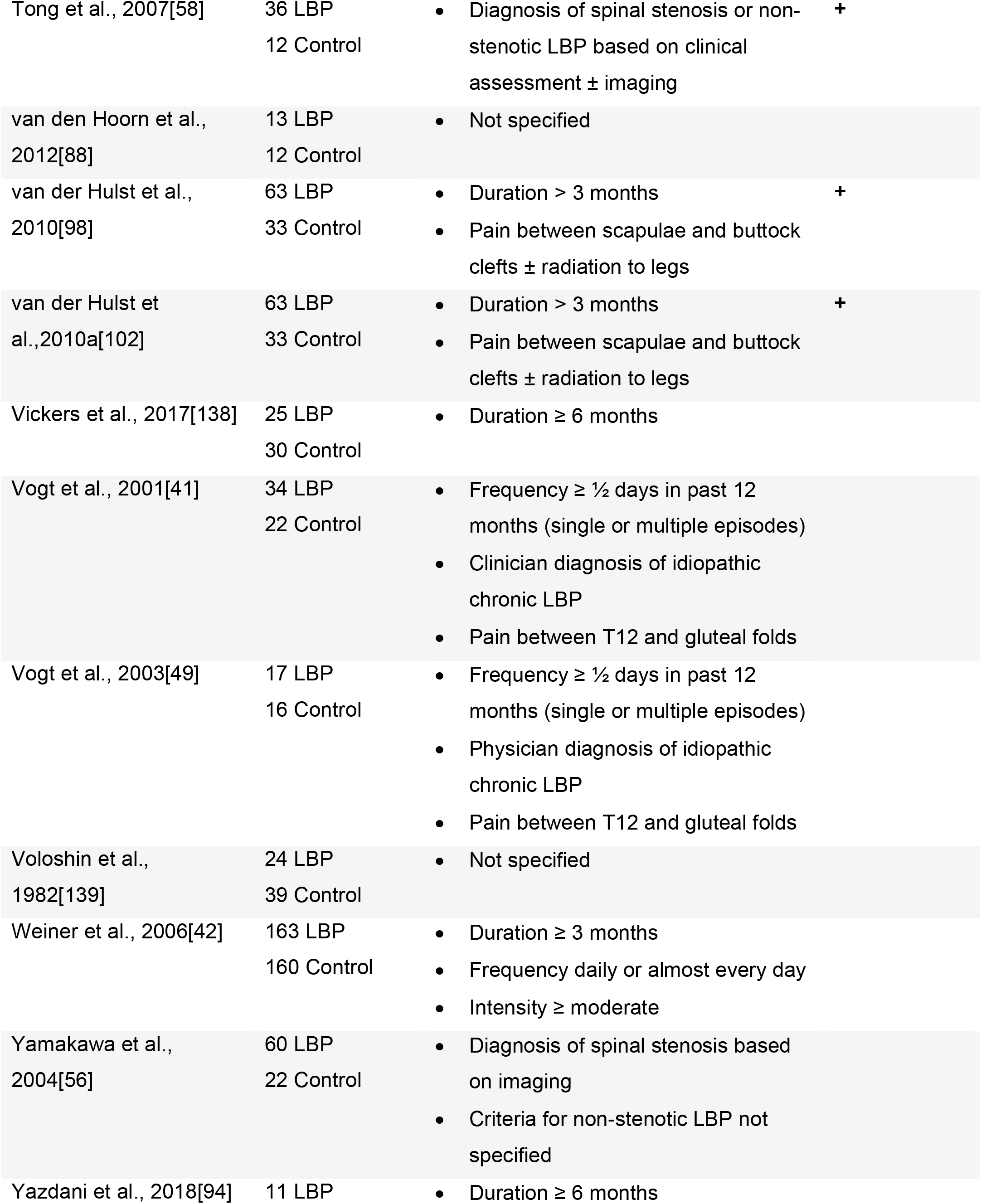
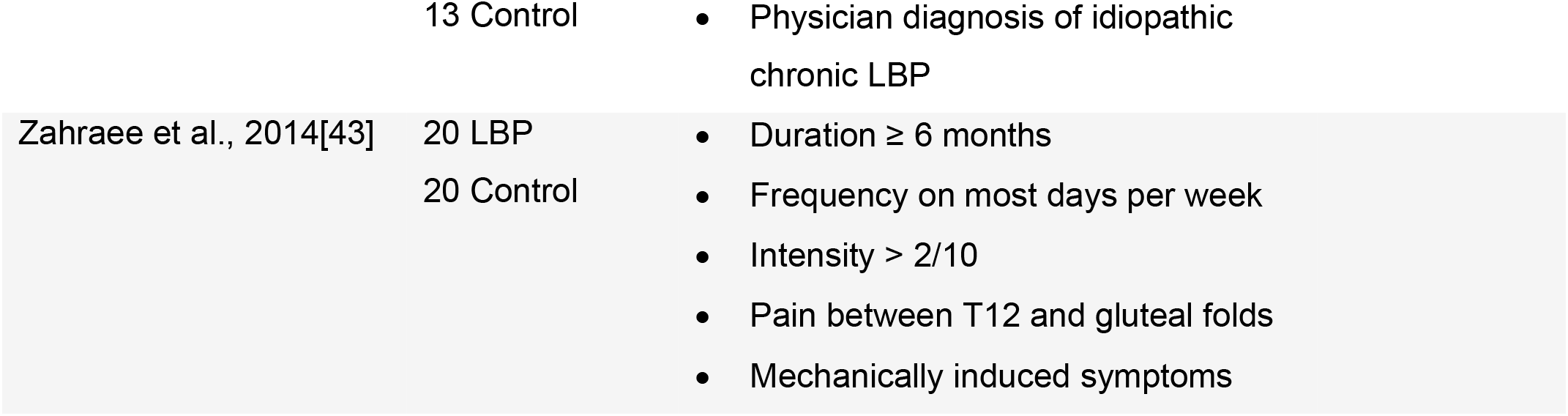
Summary of included studies

Fourteen of the studies recruited subgroups of individuals with LBP based on pathoanatomical diagnoses. Among these articles, 32 participants were diagnosed with lumbar disc herniation[50–54]. One hundred and eight participants were described as having spinal stenosis[55–58]. Forty participants had LBP with pain referred to the lower limbs[59,60]. Thirty-nine participants were described as having radiculopathy[45,46]. Lastly, one article recruited a pain group with LBP diagnosed as stenosis, degenerative instability, or disc herniation[61]. For the control groups, most studies required controls to be healthy, pain-free individuals, and they were frequently matched by sex and age to the experimental group.

Exclusion criteria varied widely across studies. Eighty studies explicitly excluded participants whose LBP was associated with known pathoanatomical diagnoses such as radiculopathy or if participants had a history of spinal surgery.

### Spatiotemporal characteristics

Thirty studies with a total of 1570 participants were included in the pooled analysis of preferred walking speed in individuals with persistent LBP. Moderate evidence with a moderate effect size indicated that individuals with LBP walked more slowly than back-healthy individuals (−0.59 [95% CI -0.77 to -0.42], I^2^ = 58% P < 0.001, test for overall effect P < 0.001, Figure 2). Two studies that quantified gait biomechanics across a range of controlled treadmill speeds noted that individuals with LBP were not able to maintain gait at controlled speeds of greater than approximately 1.4 m/s[62,63]. Pooled data with 687 participants indicated with strong evidence and a small effect size that individuals with LBP had shorter stride length when walking at preferred speed (−0.38 [-0.60 to -0.16], I^2^ = 45% P = 0.05, effect P < 0.001, Figure 2). Pooled evidence from 510 participants demonstrated a trend toward cadence also being reduced in individuals with LBP (−0.19 [-0.46 to 0.09], I^2^ = 53% P = 0.03, effect P = 0.18, Figure 2). There was moderate evidence from seven studies that duration of single limb support did not differ between groups (−0.17 [-0.56 to 0.23], I^2^ = 72% P = 0.001, effect P = 0.41)[44,64–68]. Five studies with a total of 385 participants reported step width, and pooled analysis showed no difference between groups, although there was a trend toward greater step width in the LBP groups, with moderate evidence (0.34 [-0.06 to 0.74], I^2^ = 72% P = 0.006, effect P = 0.10) [39,44,48,67,69]. In studies investigating distance walked in five minutes, individuals with LBP walked significantly shorter distances than healthy controls[70,71]. In studies involving running, preferred running speed did not differ between groups[31,35,72].

**Figure 2.**
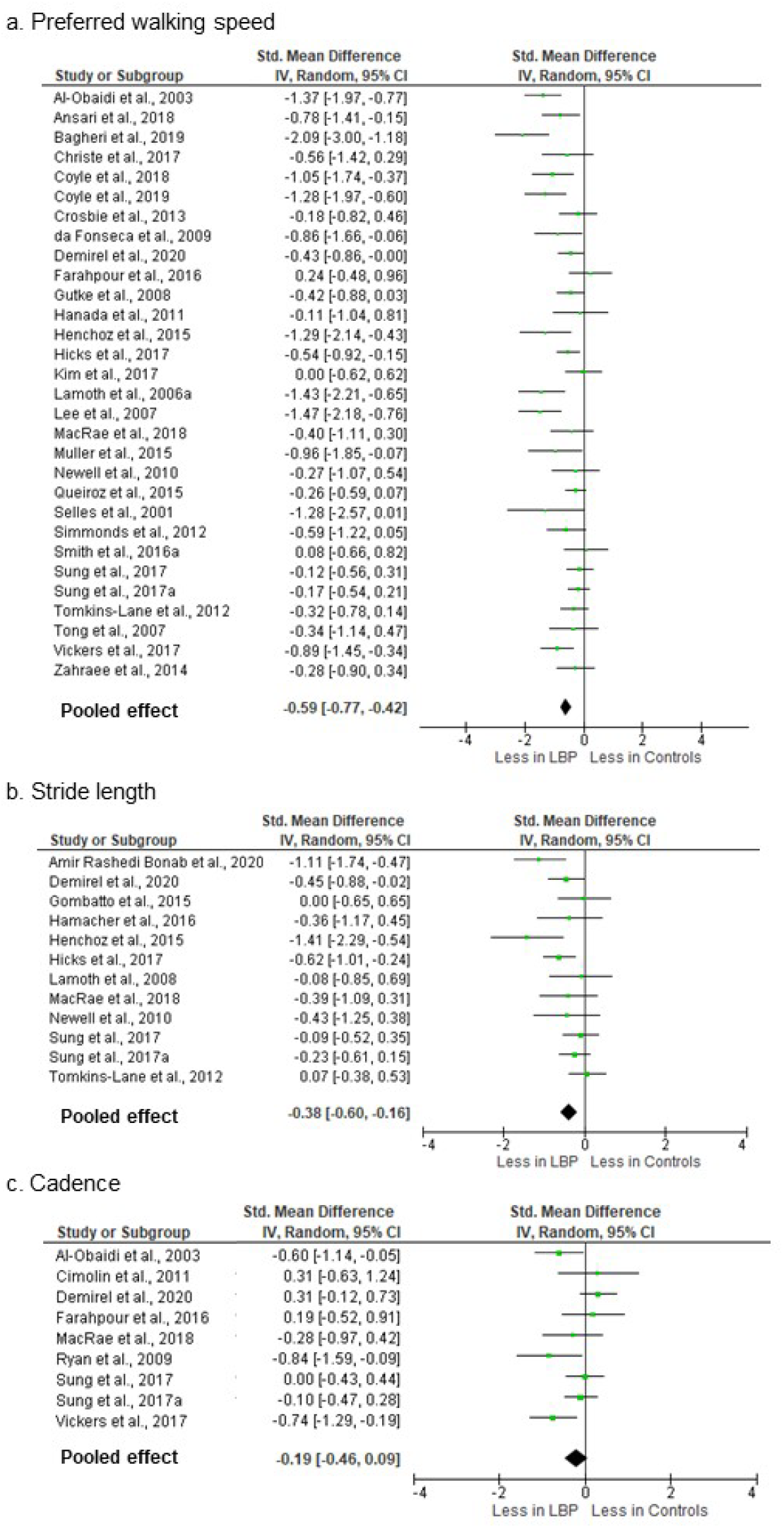
Meta-analysis of spatiotemporal gait variables. (2a) Preferred walking speed. (2b) Stride length. (2c) Cadence.

Several studies investigated how the addition of a cognitive task influenced spatiotemporal gait characteristics[38,73–75]. In individuals with LBP, the dual task condition was associated with greater or the same stride-to-stride variability of stride length [73,75].

### Kinematic characteristics - single segment/joint

Pooled analyses of five studies with 193 participants examining upper lumbar motion demonstrated with strong evidence that there was no significant difference in amplitude of motion in the axial plane in individuals with persistent LBP (0.07[-0.26 to 0.39], I^2^ = 21% P = 0.28, effect P = 0.69)[40,41,47,63,76]. These studies modeled upper lumbar kinematics with markers fixated around the spinal levels of T12[41], L1[40,47], L2[63] and L3[40,76,77]. Frontal plane upper lumbar motion was pooled from six studies and also demonstrated strong evidence that there was no difference between LBP and control groups (−0.13 [-0.39 to 0.13], I^2^ = 0% P = 0.95, effect P = 0.32)[40,41,47,63,76,77]. As few studies investigated sagittal plane lumbar motion, these data were not pooled, but no studies reported a significant difference between individuals with and without LBP[41,47,77].

Nine studies with 307 participants had axial plane data for the thorax that could be pooled[39,40,47,63,72,76,78–80]. These studies modeled upper trunk motion with markers fixated on the sternum[39,79], acromioclavicular joints[72], and/or the spinal levels of C7[39,72], T1[40,76], T3[63], and T6[47,80]. There was strong evidence of no difference between groups (−0.10 [-0.33 to 0.13], I^2^ = 0% P = 0.56, effect P = 0.40, Figure 3). Strong evidence pooled from six studies demonstrated that frontal plane motion also did not differ between groups (−0.16 [-0.45 to 0.12], I^2^ = 13% P = 0.33, effect P = 0.26)[39,40,47,63,76,78]. Of the studies investigating sagittal plane motion that could be pooled, there was moderate evidence for no significant difference between individuals with and without LBP (−0.54 [-1.30 to 0.22], I^2^ = 0% P = 0.40, effect P = 0.17)[39,47,72,78]. Intra-subject stride-to-stride variability of lumbar or thoracic kinematic motion was reported in several studies, but without consistent methodological approach or findings[41,63,81]. During running, upper trunk motion in the axial plane was reported as being less in individuals with LBP[72] or the same[32,34] and there was no difference in sagittal[32,34,72] or frontal plane motion[32,34].

**Figure 3.**
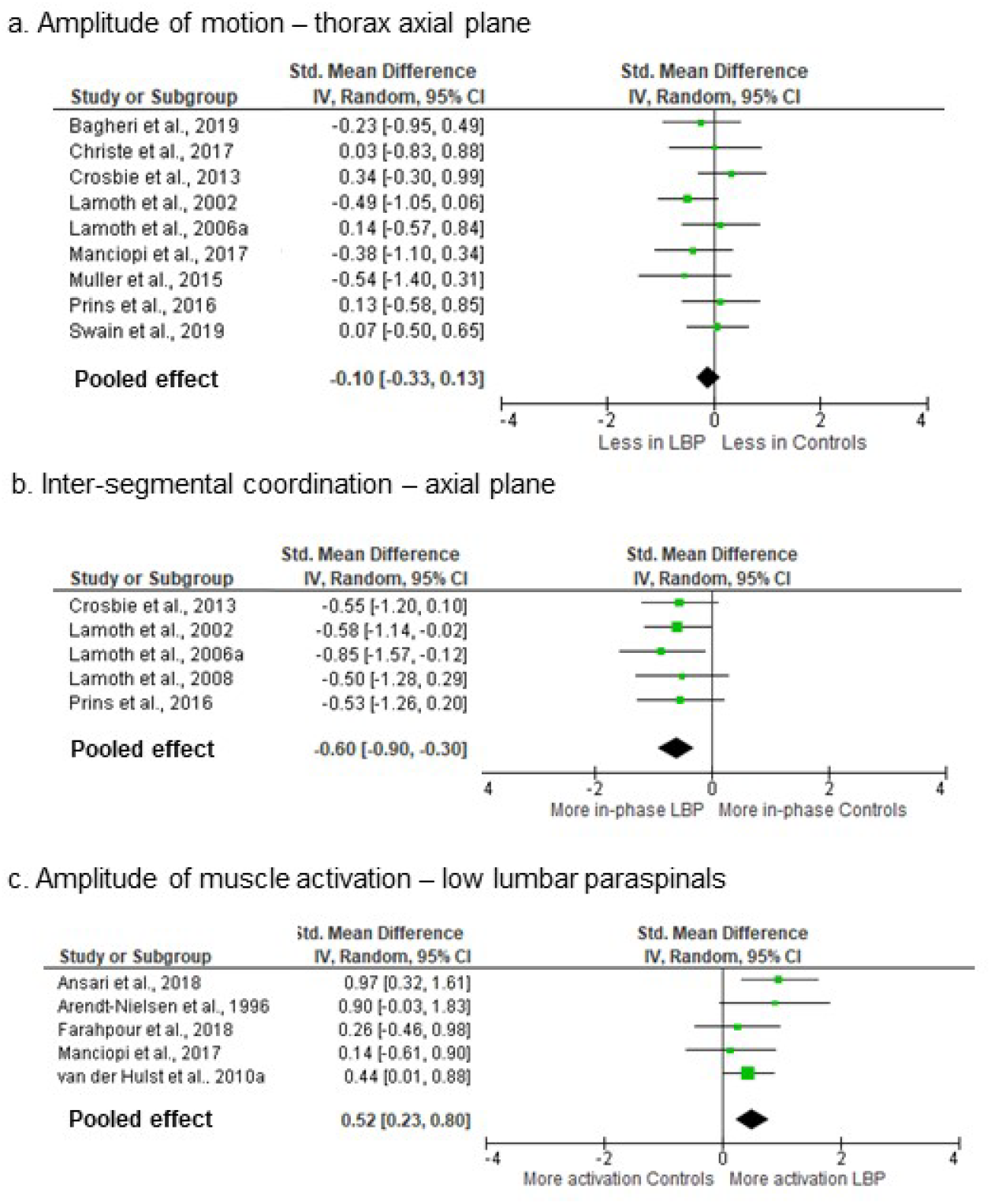
Meta-analysis of kinematic and EMG gait variables. (3a) Axial plane thoracic motion. (3b) Axial plane inter-segmental coordination. (3c) Amplitude of paraspinal activation.

The effect of performing a dual cognitive task at the same time as walking on the amplitude of lumbar or thoracic kinematics did not differ between groups[38], but did result in increased stride-to-stride variability of upper trunk motion in individuals with LBP[74].

Data from five studies with 179 participants investigating pelvis kinematics in the axial plane during walking were pooled[41,47,63,72,80]. The pelvis was modeled with markers on the sacrum[41,47,63,80] and greater trochanters[72]. Moderate evidence indicated no significant difference between groups (−0.12 [-0.42 to 0.18], I^2^ = 50% P = 0.09, effect P = 0.43). Similarly, in the frontal plane, there was strong evidence that the amplitude of pelvic motion did not differ between groups (−0.09 [-0.75 to 0.58], I^2^ = 43% P = 0.15, effect P = 0.80)[41,47,63,82]. Sagittal plane pelvis kinematics were only available in two studies and neither reported a significant group difference[41,47]. During running, one study utilizing a controlled running speed found that axial plane pelvic motion was reduced in individuals with persistent LBP[72] and another using participant preferred running speed reported that it was greater in individuals with LBP[32].

Eight studies reported hip kinematics during steady-state gait[47,49,53,83–87]. Pooled data available from four of these studies with 128 participants indicated limited evidence for no difference in total sagittal plane hip motion in individuals with LBP (−0.08 [-0.43 to 0.27], I^2^ = 0% P = 0.94, effect P = 0.65). In the frontal plane, two studies reported reduced motion[84,85], with a large effect size occurring in a study of obese adults[85] and two reported no difference[53,83]. In the axial plane, two studies reported no difference[53,83] and one reported decreased motion in individuals with LBP[84]. Knee flexion during late stance or swing phase was reduced in three out of five studies that reported knee kinematics[72,83–85,87], but there were no consistent trends evident for frontal or axial plane knee motion, or for ankle motion.

Two studies reported sagittal plane hip kinematics during running and found no difference in motion between individuals with and without LBP[30,35]. Similarly for running, sagittal plane knee motion either did not differ[34,72] or was reduced[30] in individuals with LBP and sagittal plane ankle motion was not significantly different[30,72].

### Kinematic characteristics - inter-segmental coordination

Multiple studies investigated coordination of kinematic motion between spinal segments during steady-state gait[31,32,47,63,75,79,80,88–91]. Most examined coordination in motion between the thorax and pelvis across a variety of controlled walking speeds. Time- and frequency-domain techniques were used to quantify phase relations between segments. In the axial plane, pooled analyses of 185 participants demonstrated with strong evidence and a moderate effect size that motion between the thoracic spine and the lumbar spine/pelvis was significantly more in-phase in individuals with LBP than controls (−0.60 [-0.90 to -0-.30], I^2^ = 0% P = 0.96, effect P < 0.001, Figure 3). This finding was supported by three out of four additional studies from which data could not be pooled[31,88,89,91]. Multiple studies also investigated the stride-to-stride variability of inter-segmental coordination in the axial plane during steady-state gait[31,63,75,88,90,91], with three reporting less variability in individuals with LBP[88,90,91].

Fewer studies reported frontal or sagittal plane intersegmental coordination and there was insufficient data available to pool the findings. In the frontal plane, coordination between the thorax and lumbar spine/pelvis was reported as being more in-phase in individuals with LBP[31,32,47] or the same[63]. In the sagittal plane, one study reported more in-phase coordination in individuals with LBP[92]and two others sharing the same cohort reported no difference [31,32]. Stride-to-stride variability in sagittal thorax-pelvis coordination was reported as being less in individuals with LBP[92] or the same[31].

### Kinetic characteristics

Five studies reported kinetic measures in the lower extremities during walking[83,85,86,93,94]. In three studies examining sagittal plane total net joint moments at the hip, there were no differences between individuals with and without LBP[83,86,93]. During running, sagittal plane hip moments did not differ between groups[30,35]. One running study reported increased external flexion moment at the knee in individuals with LBP[35] but there was no difference in another study[30].

### Ground reaction forces

Five studies with 138 participants investigated ground reaction forces during gait[43,59,62,65,94]. Limited evidence from pooled data with high heterogeneity suggested that there was no difference between groups in peak vertical ground reaction forces during either the first (0.29 [-0.54 to 1.11], I^2^ = 82% P < 0.001, effect P = 0.49) or second vertical force peaks (−0.21[-0.86 to 0.46], I^2^ = 72% P = 0.01, effect P = 0.56).

### Muscle activation characteristics

Fifteen studies included EMG measures of the paraspinal and abdominal musculature during walking, using preferred and controlled walking speeds[36,39,49,61,63,83,90,95–102]. Of these, fourteen used surface EMG electrodes and one used intramuscular EMG. Most studies investigated amplitude of muscle activation, with two investigating timing of activation[36,49]. During steady-state gait, pooled analyses of data from 210 participants for surface EMG of the low lumbar paraspinal musculature across the entire stride cycle or within the stance phase indicated with moderate evidence and a moderate effect size that individuals with LBP had greater amplitude of activation (0.52 [0.23 to 0.80], I^2^ = 1% P = 0.40, effect P < 0.001, Figure 3).

For the abdominal musculature, two high quality studies reported increased rectus abdominis activity during some or all phases of gait[95,98], one reported decreased activity[96] and two studies, including one high quality study, reported no difference in activity between groups[83,97]. Three studies reported no difference in amplitude of external oblique activation[83,97,98] and one high quality study reported increased activation during some subphases of gait[95]. One study reported increased peak activation in internal oblique compared with controls[83], one reported variable results depending on subphases of gait[96] and another reported decreased activity during several gait subphases[97].

## DISCUSSION

Individuals with persistent LBP walk differently than back-healthy controls. These differences are most evident in spatiotemporal characteristics, in patterns of inter-segmental coordination, and in paraspinal muscle activation. This is despite the fact that walking is often pain-relieving in individuals with acute or persistent LBP[28,63], and is often recommended as part of a rehabilitation program[11,103]. Current evidence does not indicate that LBP is associated with a difference in the amplitude of motion in the trunk or lower extremities during walking or running. To our knowledge this is the first comprehensive systematic review and meta-analysis of walking and running gait in individuals with LBP.

Pooled data demonstrated that individuals with persistent LBP choose to walk more slowly than individuals without back pain. In two studies that reported that LBP patients were unable to walk at controlled fast speeds[62,63], it was unclear if this inability was due to pain, fear of pain, or deconditioning. Al Obaidi et al.,[64] examined influences on preferred walking speed and found that fear avoidance and pain anticipation significantly predicted reduced walking speed in individuals with persistent LBP. It is possible that individuals with LBP use a strategy of slower walking velocity, reduced stride length, and decreased cadence to minimize the kinematic and kinetic demands of walking[28,59]. Unfortunately, as very few studies quantified spatiotemporal gait characteristics at controlled gait velocities, or adjusted for gait velocity in their analyses[48,63,77], there is insufficient evidence to separate the influence of slower gait velocity from the independent effects of LBP on these characteristics. Clinically it may be important to assess gait at a range of speeds in individuals with LBP to identify mechanisms underlying reduced speed and impaired stride length and cadence.

This study found strong evidence for altered phase relations between motion in the thorax and the pelvis during walking in individuals with persistent LBP. In back-healthy controls, the pattern of coordination, or relative motion, between the upper trunk and pelvis in the axial plane is speed dependent, becoming more anti-phase as speed increases[63]. Even when walking at controlled speeds, individuals with LBP exhibited greater in-phase movement patterns. This may be due to reduced ability to dissociate movement between the trunk and pelvis in these individuals. As anti-phase coordination during fast walking helps to generate elastic recoil between the thorax and the pelvis and may also contribute to minimizing total body angular momentum in the axial plane[104], the reduction in anti-phase coordination in individuals with LBP may contribute to decreased gait speed and reduced stride length.

Our meta-analysis demonstrates that individuals with LBP have greater lumbar paraspinal activation during walking. Phasic muscle activity in the paraspinals occurs bilaterally at initial contact and during the double support phases of the gait cycle[105,106]. This activation controls sagittal and frontal plane motion between the trunk and the pelvis[107]. The amplitude of this activity is low, typically less than 20% of maximum voluntary activation for walking[36,108] although this increases to up to 100% of maximum for fast running[108]. Acutely, increased activation during gait may be adaptive if it serves to reduce motion and project pain-sensitive tissues. After the acute phase, it may also be a compensation for the muscle weakness related to atrophy and fatty infiltration that occurs in multifidus in response to back pain[109] or for proprioceptive dysfunction[110,111]. However, over time this increased activation in individuals with LBP may contribute to recurrence due to increased compressive spinal loading[112]. Increased paraspinal activation may also result in increased axial stiffness between the upper trunk and the pelvis, partially explaining the reduced trunk/pelvis dissociation described above[88,104].

In comparison with the paraspinals, abdominal muscle activity during locomotion is much more variable between individuals and more dependent upon locomotor speed[105,107,108]. This variability within healthy individuals is perhaps due to the redundancy of the abdominal muscle system and likely accounts for the lack of consistent differences in abdominal activation in individuals with LBP in the present review. It should be noted that all but one of the studies in this review used surface EMG. Surface EMG cannot selectively quantify activation in the transversus abdominis and multifidus muscles[113,114]. Isolated postural impairment of these deep muscles has been a focus of LBP research and treatment for some years. However, our findings are consistent with a recent systematic review of anticipatory postural adjustments indicating that the postural function of the superficial muscles in the abdominal and paraspinal systems are also affected by LBP[115].

Current evidence does not consistently demonstrate a significant difference in joint or segmental excursion in the trunk or lower extremities during walking or running in individuals with LBP. For the thoracic and lumbar spines, joint range of motion utilized during walking and slow running is a small proportion of the available range[108]. This is in contrast with other activities such as standing forward flexion where significant reductions in lumbar range of motion in individuals with LBP have been observed[116]. The amplitude of hip and knee motion during walking and running is a greater proportion or available range, but the current evidence does not consistently support interdependency between back pain and lower limb gait kinematics.

### Limitations

The limited number of available studies that investigate running precluded meta-analyses of the running biomechanics in individuals with LBP. As noted earlier, many studies quantified walking biomechanics at participants’ preferred walking speeds. This makes it difficult to determine if the observed impairments in characteristics like stride length in individuals with LBP are evident even when walking speed is controlled. Additionally, in this review we were unable to probe differences in gait between sub-groups of individuals with LBP. The inconsistent sub-grouping or classification of individuals with persistent LBP remains problematic. Multiple classification systems based on biomechanical or kinesiopathological factors have been proposed, but none are fully supported by available evidence[117]. Several studies in this review recruited participants based on pathoanatomical diagnoses such as herniated lumbar discs, degenerative instability, or spinal stenosis. Studies varied in how these pathoanatomical diagnoses were made, and it is now widely recognized that pathoanatomical findings do not adequately inform clinical presentation or outcome[118]. Some studies investigated patient sub-groupings based on age, sex, weight, pain severity, or psychosocial factors. However, the heterogeneity of the participants included in this review results in greater generalizability of the findings to the broader clinical population.

## CONCLUSION

We found that individuals with LBP exhibit different biomechanical characteristics during gait than back-healthy controls. Differences are most evident in spatiotemporal characteristics, thorax/pelvis coordination, and paraspinal muscle activation. However, it is not known if the strategies evident in individuals with LBP during gait are adaptive or maladaptive. Prospective research following the transition from acute to persistent pain or symptom resolution will provide insight into the effect of these altered gait mechanics on the trajectory of back pain symptoms over time.

## Supporting information

Prisma checklist

Search strategy

## Data Availability

All data generated or analyzed during this study are included in this published article [and its supplementary information files

## Acknowledgements

The authors would like to thank Ivan Portillo MLIS AHIP for his assistance in refining the search strategy and search terms.

