## Supplementary material for "Do people with low back pain walk differently? A systematic review and meta-analysis": Search strategy

Supplementary information

CINAHL

(walk or walking or run or running or locomotion or gait or perturb or perturbation or turn or turning ) AND ( lower back pain or low back pain or low backache or lumbar pain or lumbago or nonspecific low back pain ) NOT ( cancer or surgery or post surgery ) NOT ( animal testing or animal experimentation or animal research or laboratory animals or animal or animals) NOT ( amputation or amputee or amputees or limb loss )

PubMed

(((("walk"[All Fields] OR "walking"[All Fields] OR "run"[All Fields] OR "running"[All Fields] OR "locomotion"[All Fields] OR "gait"[All Fields] OR "Gait"[All Fields] OR "perturb"[All Fields]) AND ("lower back pain"[All Fields] OR "low back pain"[All Fields] OR "low-back pain"[All Fields] OR "low backache"[All Fields] OR "lumbar pain"[All Fields] OR "lumbago"[All Fields] OR "nonspecific low back pain"[All Fields])) NOT ("cancer"[All Fields] OR "surgery"[All Fields] OR "post surgery"[All Fields])) NOT ("amputations"[All Fields] OR "amputation"[All Fields] OR "amputee"[All Fields] OR "limb loss"[All Fields])) NOT ("animals"[MeSH Terms:noexp] OR ("animals"[MeSH Terms:noexp] OR ("animals"[MeSH Terms:noexp] OR ("animals"[MeSH Terms:noexp] OR ("animals"[MeSH Terms:noexp] OR ("animals"[MeSH Terms:noexp] OR ("animals"[MeSH Terms:noexp] OR ("animals"[MeSH Terms:noexp] OR ("animals"[MeSH Terms:noexp] OR animal[All Fields]))))))))) AND "humans"[MeSH Terms]
